# Contact bioassay in tandem with HPLC analysis of the active ingredients in LLINs revealed hidden possible causes of the unabated rise in malaria cases

**DOI:** 10.1101/2022.11.07.22282036

**Authors:** Michael O. Kusimo, Sulaiman S. Ibrahim, Nelly O. Kusimo, Alison O. Nwokeoji, Omoniyi K. Yemitan

**Affiliations:** Centre for Research in Infectious Diseases (CRID), P.O. Box 13591, Yaoundé, Cameroon; Vector Biology Department, Liverpool School of Tropical Medicine (LSTM), Liverpool L3 5QA, UK; Department of Biochemistry, Bayero University, PMB 3011 Kano, Nigeria; Society Empowerment for Transformation Initiative (SETI), 10 Chief Ogbonda Street Artillery, Rumukurushi, Port-Harcourt, Rivers State, Nigeria; Dept. of Chemical and Biological Engineering, University of Sheffield, Mappin Street, Sheffield S1 3JD, UK; Department of Pharmacology, Therapeutics and Toxicology, Lagos State University College of Medicine, Ikeja, Lagos, Nigeria

**Keywords:** Malaria elimination, HPLC, bed net, environmental pollution, Nigeria

## Abstract

**Background:** The World Malaria Report 2021 revealed that Nigeria accounted for 27% of 241 million malaria cases worldwide and 31% of 602,000 malaria deaths. This WHO report is in sharp contrast to the ambitious goal of the National Malaria Strategic Plan (NMSP) 2014-2020, which aimed to transition Nigeria from malaria control to malaria elimination status by 2020. In this study, we combined contact bioassay with HPLC analysis to investigate how the end users’ treatment of Long-Lasting Insecticide Treated Nets (LLINs) inadvertently contributes to the unabated rise in malaria cases.

**Methods:** We randomly selected a few LLINs used under normal household conditions in Port-Harcourt and one on sale in Lagos, Nigeria. The continued potency of these LLINs to protect the users against malaria vectors, assuming the local vectors are insecticide-susceptible, was evaluated by exposing laboratory-susceptible *Anopheles gambia* Kisumu strain to the nets according to the WHO cone bioassay protocol. After the exposure, the active ingredients (AI) in the LLINs were extracted and analysed using reverse phase HPLC to establish the quantity of the AIs and correlate it to bio-efficacy. The AI loss per wash was also computed based on the information provided by the users.

**Results:** The labels on the LLINs revealed them to be Royal Sentry, PermaNet 2.0 and DuraNet. The bio-efficacy of these LLINs under household usage is less than two years. AI lost per wash was higher by 2.5-fold in LLIN made of polyethylene nets than polyester nets: 0.35g/Kg/wash and 0.14g/Kg/wash, respectively. The AI lost per wash increased by 31% for net exposed longer to the harsh environmental conditions of this region. The LLIN (ParmaNet) purchased in the open market has the highest AI concentration but only achieved 70.62% mortality, far below the acceptable standard.

**Conclusion:** The AI in the LLIN bought from the open market was the highest, but of poor quality. This bed net was a counterfeit product, not approved by WHOPES. More importantly, this study revealed that field examination of LLINs under normal household use should be examined per region for policy makers to have informed knowledge of the duration of the bio-efficacy of the LLINs being distributed.

## Background

The successes achieved in malaria control in the past 20 years have been possible through, among other factors, the introduction of Insecticide Treated Nets (ITNs) and Indoor Residual Spraying (IRS) [1]. The bed nets provided the protective barrier preventing mosquito bites, and the insecticides coated on the bed nets killed the malaria vectors, thus reducing the malaria burden and saving lives [2]. However, the rigour of retreating ITNs with insecticides after frequent washes and the related cost [3, 4] led to the development of more robust Long Lasting Insecticide Nets (LLINs) [5]. This new generation of insecticide-treated bed nets does not need re-treatment and remains active after 20 washes or three years of using them [6]. Among the first generation of LLINs approved by the World Health Organization Pesticide Evaluation Scheme (WHOPES) are PermaNet and Olyset [4, 7], containing pyrethroid insecticides: deltamethrin and permethrin, respectively. Conversely, due in part to the use of pyrethroid as the only insecticide for the production of LLINs, malaria vectors are increasingly developing resistance to these LLINs reversing the gains achieved in stemming the malaria burden across the endemic regions, especially in sub-Sahara Africa [8, 9].

Resistance to pyrethroids by mosquitoes has been widely documented and is based on the development of four central resistance mechanisms by these disease vectors [10]. The vectors could thicken/vary the composition of their cuticles, thus preventing the absorption of the insecticides [11], or avoid getting close to the insecticide altogether [12]. Better still, they could alter the binding sites of the insecticides [13-15] or overexpress multiple detoxification enzymes to neutralize and eliminate the insecticides absorbed into their bodies [16, 17]. One of the most effective detoxification enzyme groups is the P450 enzymes, a phase 1 detoxification enzyme that enhances the solubility of pyrethroids for elimination once absorbed by the vectors [18]. Mosquitoes have over 100 P450 genes that could be deployed to degrade insecticides, and many have been functionally validated to be pyrethroid metabolizers [19]. The vectors’ development of resistance against pyrethroids warranted the development of next-generation LLINs fortified with an inhibitor of the P450s, piperonyl butoxide (PBO) or pyriproxyfen, a mimic of juvenile hormone, to revert the efficacy of pyrethroids [20]. Notwithstanding the development of resistance by malaria vectors to LLINs, insecticide-treated bednet is still a preferred vector control tool. It has proven to continue to protect lives when used than not being used at all [21].

From 2001 to 2014, the Nigerian government launched four malaria campaigns to control the country’s malaria epidemic [22]. The most recent National Malaria Strategic Plans (NMSP) aimed to transition the country to pre-elimination status and reduce malaria-related death to zero by 2020 [23]. However, the effects of these set goals are not manifesting because Nigeria has consistently topped the list of malaria-burdened countries since then [24]. The current World Malaria Report 2021, for the operational year 2020, revealed that Nigeria accounted for 27% of 241 million malaria cases worldwide and 27% 602, 000 of all global malaria deaths [25]. One of the critical strategic plans of NMSP was to achieve universal coverage of 1:2 (net: person) LLINs. However, according to a recent study carried out in rural communities of River State, the range was 1 to 4 instead of 1:2 and more worrisome is that over 90% of those that have the LLINs do not usually sleep under them because of discomfort and nightmares they claim to experience when under the protection of the bed nets [26].

In this study, we randomly selected three households in Port-Harcourt, the capital city of Rivers State in the south-south region of Nigeria. A fourth LLIN was purchased in a store in Lagos, Nigeria. The WHO cone bioassay was carried out on the LLINs using the *Anopheles gambiae* lab susceptible Kisumu colony. After the cone assays, the active insecticide (AI) in the LLINs was extracted and quantified using reverse phase HPLC analysis to confirm the content and quantity of the AIs remaining in the LLINs after several washes by these households.

## Methods

### Collection and Identification of the LLINs

LLINs were randomly collected in 2019 from three households that indicated from a previous study that they own and sleep under LLINs in Port-Harcourt, Nigeria. The LLINs were identified through their product labels on the net. The age of the LLINs, frequency of wash, household size, and number of people that slept under the LLINs and the source of the LLINs were documented. We gave each household a new Permanent 2.0 to replace the LLINs collected. We also examined a new LLIN purchased from a popular pharmaceutical shop in Lagos, the capital city of Lagos State, South-West of Nigeria. A fifth insecticide-free net was used as control.

### Evaluation of physical integrity of the LLINs

The LLINs collected from the households were examined for holes, seam integrity, and evidence of repairs to establish the physical conditions of the LLINs following previously established protocol [27].

### Determination of the bio-efficacy of the LLINs

To assess the effectiveness of the insecticides embedded in the LLINs against malaria vectors, we carried out a cone bioassay following WHO protocol [28]. Two fragments of 25 cm by 25 cm, one from the side and the other from the roof, were cut out from each LLIN. Five unfed 3-5 day old *A. gambiae* laboratory-susceptible Kisumu strain was introduced into the cone. Five sets of cones with mosquitoes were exposed to five positions on each LLIN fragment for 3 mins. A total of 10 positions and 50 mosquitoes per LLIN were tested. After exposure, the mosquitoes were transferred into paper cups and provided with a 10 % sugar solution. Knockdown (KD) was recorded after one hour of exposure and mortality after 24 hours. All these operations were carried out at a controlled temperature of 23 °C and humidity of 65 % in the insectary laboratory of the Centre for Research in Infectious Diseases (CRID), Yaounde, Cameroon.

### HPLC analysis of the amount of active ingredients in the LLINs

A smaller fragment of 10 cm by 10 cm was cut out from each of the pieces of the LLINs used for the cone assay, and three more fragments were also cut out from other parts of the main nets to have five replicates for the AI quantification per bed net. These pieces were weighed, chopped, and placed in a glass vial. Acetone was used to extract AIs in three fragment replicates by adding 10 mL acetone, vortexing for 5 min and sonication for 5 min. Acetonitrile was used for the remaining two replicates, adding 10 mL and vortexing for 5 min without sonication. One millilitre from the acetone extractions was placed in a new vial, evaporated and re-suspended in 1 mL acetonitrile. The acetonitrile extraction was used directly. We further examined if all the AIs in the net were fully extracted by repeating the extraction process. The extracted solution was decanted and the net rinsed with acetonitrile twice before repeating the process. About 200 µL of all the extractions were filtered using 4mm SYR filter PTFE, 0.2 µM into HPLC vials and 10 µL injected into the HPLC machine. Isocratic mobile phase of ACN: H^2^O (80:20) was used at 1 mL per min Acclaim™ 120 C18 5 µm column, and analytes were detected at UV of 226 nm. Insecticide standards of deltamethrin and alpha-cypermethrin were prepared, and the level of detection (LOD) and level of quantification (LOQ) of the insecticides on the HPLC machine were determined using the linear regression method [29, 30].

## Results

### Details of the LLINs being used under normal household conditions

Routine assessment of the continued potency of bed nets distributed in the field is important to monitor their effect on malaria control after being used under normal household conditions, which could have adverse effect on their durability. The earlier bed nets that have lost their bio-efficacy are withdrawn from the field, the quicker it will be to prevent the local malaria vectors from developing resistance to the insecticides in the bed nets. Here using bed nets in use in three households, we describe a simple workflow for effectively monitoring potency of bed nets collected from the field by correlating the quality of LLINs, irrespective of age or quantity of the insecticides in the net, to bio-efficacy.

Household 1 (H 1) has been using Royal Sentry LLIN, collected from the hospital, for two years, and washing was done once in 2 months with soap and water. The label on the LLIN shows it was one of those distributed by NMEP (National Malaria Elimination Program). It has the National Agency for Food and Drug Administration and Control’s (NAFDAC) number with the logo of the coat of arms of the federal republic of Nigeria. This LLIN was manufactured in December 2015 and was set to expire in November 2020. There are three individuals in this household, but only the mother and the child sleep under the LLIN. Household 2 (H 2), a family of 5, received their LLIN from a neighbour. They have been using it for eight months and have washed it eight times. The label on the net is typically displayed on the Vestergaard website: PermaNet 2.0, 2008, as manufactured date with no expiry date. The LLIN was 12 years old as of 2020 when this experiment was done but was newly opened and in use for just eight months. It did not have the NAFDAC number. There are five individuals in this household, but only the mother and a child sleep under the LLIN. Household 3 (H 3) has been using their LLIN, a gift from a friend, for a year. The label on the LLIN shows it was one of those meant for mass distribution by the federal government of Nigeria. The DuraNet label has the logo of the coat of arms of the federal republic of Nigeria; NOT FOR SALE distributed by NMEP and a NAFDAC number. Washing was done monthly, and only one person in a family of three sleep under the LLIN. Bed net coverage for these three households did not meet the WHO universal coverage of one LLIN for two persons (1:2), Table 1.

**Table 1.** Detail of LLINs sampled from Port-Harcourt and Lagos.

|  | Household 1 | Household 2 | Household 3 | LLIN on sale |
| --- | --- | --- | --- | --- |
| Number of persons | 3 | 5 | 3 |  |
| Number of LLIN in household | 1 | 1 | 1 |  |
| LLIN coverage | 1:3 | 1:5 | 1:3 |  |
| Brand of LLIN | Royal Sentry | PermaNet 2.0 | DuraNet | PermaNet |
| Colour of LLIN | Light green | White | Dark Green | White |
| LLIN polymer type | Polyethylene | Polyester | Polyethylene | Polyester |
| Active ingredient | α-Cypermethrin | Deltamethrin | α-Cypermethrin | Deltamethrin |
| Concentration | 5.8 g/Kg (261 mg/ m<sup>2</sup>) | 1.8 g/Kg | 5.8 g/Kg (261 mg/ m<sup>2</sup>) | 1.8 g/Kg |
| NAFDAC number | A5-0641 | - | A5-0813 | 04-7508 |
| Source of LLIN | Hospital | Neighbour | Friend | Pharmaceutical store |
| How long in use | 2 years | 8 months | 1 year |  |
| Frequency of wash | Once in 2 months | Once a month | Once a month |  |
| No of wash | 12 | 8 | 12 |  |
| Label on LLIN | Yes | Yes | Yes | No |
| Mfg. date | 12/2015 | 04/2008 | 10/2016 | 04/2018 |
| Exp. Date | 11/2020 | - | 10/2021 | 04/2023 |
| How many people sleep under the net | 2 | 2 | 1 |  |
| Regularly sleep under the LLIN | Yes | Yes | No |  |

The fourth LLIN was bought from a pharmaceutical store in Lagos. PermaNet is the only LLIN type being sold by a few of the stores visited. The LLIN bought has a NAFDAC number, manufactured and expiring dates of 04/2018 and 04/2022, respectively, stated on the product package. It has no product label, which is unusual of Vestergaard products.

### Physical evaluation of the LLINs

All the LLINs from the households were in good condition. No hole and no sign of repair was observed in any of them.

### Bio-efficacy of the LLINs

The cone bioassay performed on these LLINs using the lab-susceptible *An. gambiae* Kisumu strain revealed that all three LLINs being used by the households and those bought in Lagos were able to knock down the susceptible mosquitoes after 3 min exposure with none below 95% KD after 1 hr exposure, Figure 1. However, only PermaNet 2.0 from H 2 and DuraNet from H 3 attained above 80% mortality benchmark for cone bioassay achieving 91.79 and 98% mortality, respectively. The Royal Sentry from H 1 has been used for two years and washed 12 times. Nevertheless, these conditions are still within the three years and 20 washes stipulated by WHOPES [28]. DuraNet from H 3, though only in use for a year, had also undergone 12 washes and is still active. Both are polyethylene net and coated with α-cypermethrin.

**Figure 1:**
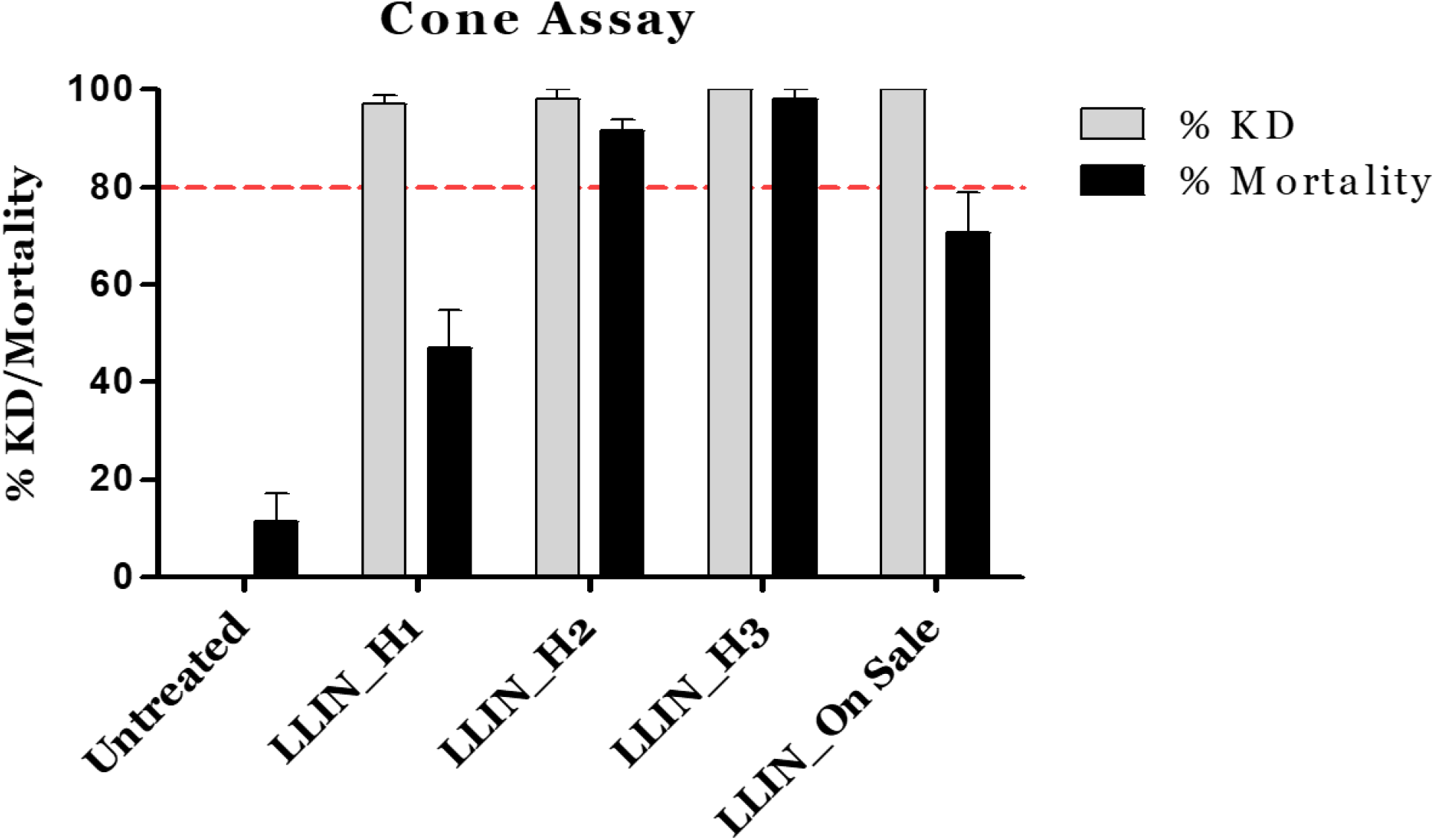
Residual bio-efficacies of the LLINs against susceptible *An. gambiae* Kisumu strain. LLIN being used by H1 has lost its bio-efficacy, and LLIN on sale in Lagos did not meet the WHOPES mortality benchmark.

The PermaNet in circulation in Lagos gave a worrisome result. The new LLIN achieved only 70.62% mortality. This result only confirmed our suspicion of the LLIN since Vestergaard does not market PermaNet but PermaNet 2.0 and PermaNet 3.0.

### Quantification of AI in the LLINs

The extraction of the AI with acetone or acetonitrile provided similar results, and sonicating the sample after vortexing did not have effect on the quantity extracted. We also did not detect any residual AI when we repeated extraction on the bed nets examined.

We established the limit of detection (LOD) and limit of quantification (LOQ) of α-cypermethrin and deltamethrin on the HPLC, as shown in Figure 2 computed using linear regression [29, 30]. The HPLC machine detected both type 2 pyrethroids to 7µg and quantified them up to 22 µg.

**Figure 2:**
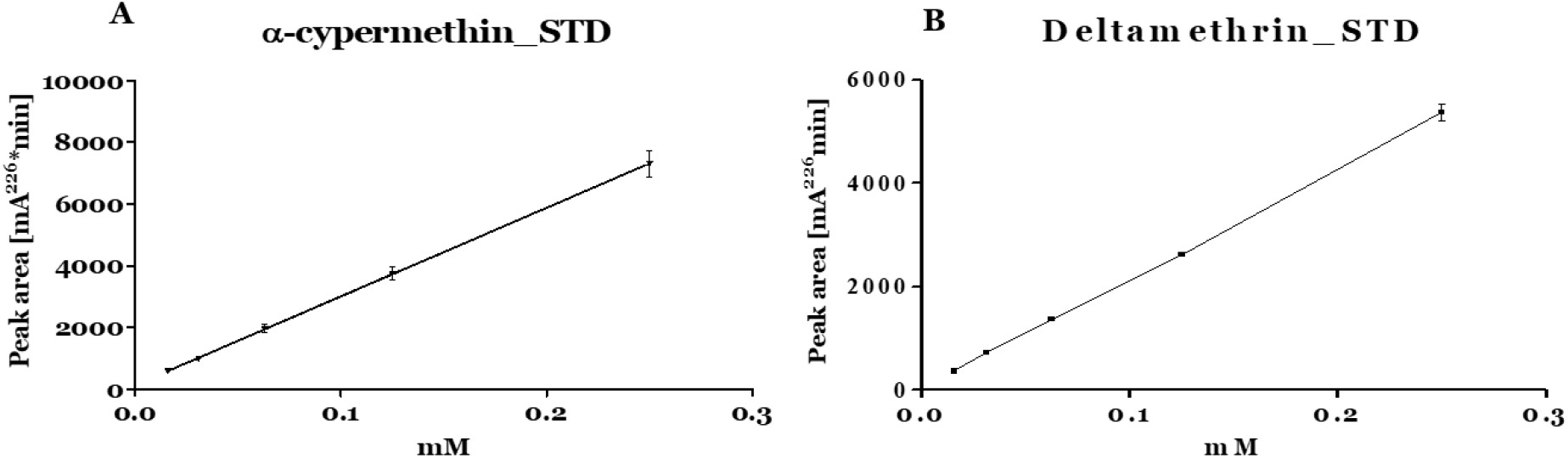
Linear regression of standard curves of α-cypermethrin and deltamethrin.

The HPLC analysis presented in Table 2 shows that 12 washes of the bed net from H 1 had decreased the AI in the LLIN by 95%, eight washes reduced AI being used in H 2 by 62.7% and AI in the LLIN from H 3 reduced by 73% after 12 washes. LLINs from H1 and H3 are polyethylene nets. AI lost per wash is higher in polyethylene nets than in polyester nets. The amount of AI quantified from the LLINs is directly proportional to the mortality observed with the cone assays. LLIN, with the lowest mortality of the susceptible mosquitoes, had the lowest concentration of AI.

**Table 2:**
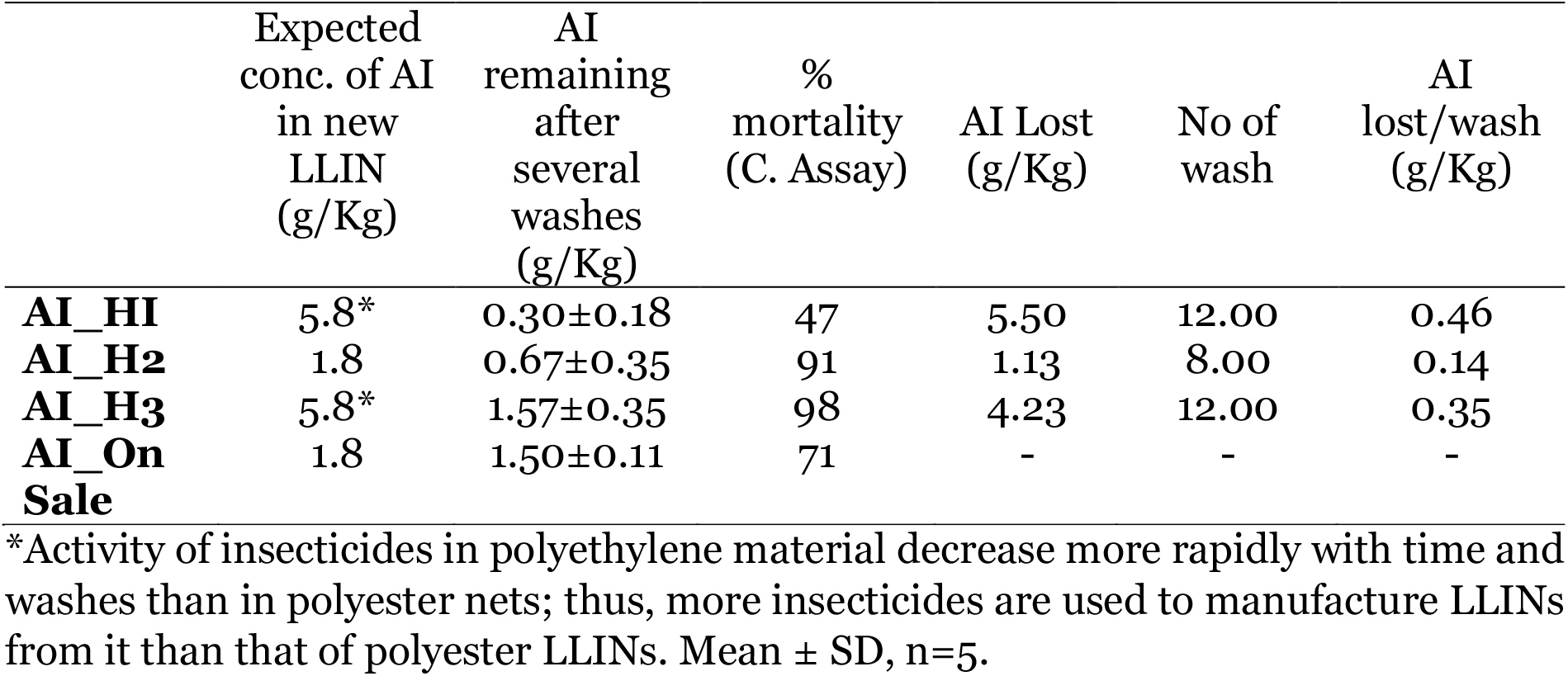
Quantities of AI remaining in LLINs and rate of lost per wash

*Activity of insecticides in polyethylene material decrease more rapidly with time and washes than in polyester nets; thus, more insecticides are used to manufacture LLINs from it than that of polyester LLINs. Mean ± SD, n=5.

Both the LLINs from H1 and H3 had been washed 12 times. However, LLIN from H1 was exposed to environmental factors for two years, whereas LLIN from H 3 was exposed for just one year. The higher AI loss in LLIN from H1 suggests the impact of environmental conditions and long usage on the insecticidal contents and efficacy of bed nets. Four more washes of LLIN in H3 at the rate of 0.35g/kg lost per wash will make it non-effective, and two more washes of H2 will take it to the level of H1.

These results suggest that the bio-efficacy of LLINs under this environmental pollution is less than two years.

## Discussion

Stamping out malaria in Nigeria will require an unwavering commitment from the government and the Nigerian people. The commitment of the Algerian government and her people in stamping out malaria is a lesson for all, as the country was declared malaria-free in 2019 [31]. The malaria parasite was first identified in this country more than a century ago [32, 33] and has been under the malaria burden ever since. It took over a decade of the consistent, unwavering fight to control and eliminate it from the country. This country invested in universal healthcare, free malaria diagnosis, well-trained health personnel, quick response to outbreaks and improved surveillance to stamp out malaria [33]. The strategies deployed by the Nigerian government through the NMSP to end the malaria burden by 2020 are useful malaria control tools that have proven effective. However, the government needs to do more in monitoring the quality of the control tools and adequately implement these malaria control strategies [34].

This study revealed the importance of evaluating the qualities of malaria control tools being used to eliminate the disease in endemic countries like Nigeria. Substandard LLINs or those that have lost their bio-efficacies while in use will increase the risk of insecticide resistance [34]. Expectedly, LLIN that give discomfort will lead to people not using it, negating the strategic plan and purpose for which the LLIN is being distributed and used as a malaria control tool [26, 35]. Regulation of the qualities of the vector control tools being used and distributed is essential so that substandard products are not allowed in the country. Production and distribution of ITN and LLINs are highly regulated by WHOPES [6]. The products undergo safety assessment which investigates the risk to humans sleeping under the LLIN; bio-efficacy of the insecticide against malaria vectors and wash resistances are evaluated [28]. Only LLINs that meet these rigorous assessments are approved and recommended by WHOPES to be used as vector control tools. These approved LLINs will ensure that susceptible mosquitoes are effectively killed, thus preventing the development of resistance mechanisms and ending the malaria burden quickly.

The discovery by this study of a counterfeit LLIN being sold in Lagos revealed further some of the hidden causes of failure to eradicate malaria cases in Nigeria. PermaNet was the first generation LLIN produced by Vestergaard submitted for quality control check with WHOPES. This LLIN failed to maintain the targeted 80% mortality [7, 36].

Vestergaard then developed a superior technology of impregnation to produce Permanet 2.0 during the process and substituted it for PermaNet. The two have the same concentration of active insecticide and polymer. The only difference is the releasing rate of the AI perfected to produce the 2.0 version [7, 36]. PermaNet 3.0 is one of the next generation LLINs adding PBO with deltamethrin [20]. The PermaNet LLIN has a choking smell that could cause discomfort if people sleep under it. Poor sleep could lead to nightmares [37]. The issue of fake and counterfeit LLINs has been going on since the beginning of the Nigerian government’s effort to control malaria in Nigeria. Table S.I 1 shows other fake LLINs previously reported and sold to unsuspecting Nigerians with phoney NAFDAC registration numbers or those confirmed for unrelated products [38].

Eliminating malaria has a substantial economic advantage for Nigeria. For instance $424.4 million was spent on malaria in 2016, out of which the government contributed 19.2% [39]. According to a WHO report, a 10% reduction in malaria incidence in high-burden and low-income countries like Nigeria is associated with an average rise of nearly 2% in GDP per capita and faster GDP growth [40].

## Conclusion

Based on the dismal progress in controlling the malaria burden, it is inevitable that the Nigerian government should start strategizing for the fifth malaria campaign. The new drive should ensure reorientation of people’s perceptions and defuse the “myth” of having nightmares when sleeping under LLINs. This is important for the LLINs being distributed to be used appropriately by the recipients, especially the vulnerable population: pregnant and nursing mothers. The health officials should be held accountable for every LLIN distributed so the bed nets get to the beneficiaries on time. The government should do more to fund the universal coverage of LLINs, and old nets should be replaced, especially those quickly losing their bio-efficacy due to the environmental pollution in the South-South oil-rich region. NAFDAC should be more responsible for adequately regulating the LLINs in the open market. They should ensure that only WHOPES-approved products are registered to be sold in Nigeria and use their regulatory powers to prosecute those using fake NAFDAC numbers to circulate substandard products.

## Supporting information

https://drive.google.com/file/d/1vGpampD07czRTXnKvfzdH4qBWXIfqpz0/view?usp=share_link

## Data Availability

All data produced in the present work are contained in the manuscript

## Abbreviations

*NMSP*: National Malaria Strategic Plan
LLINs: Long-Lasting Insecticide Treated Nets
*IRS*: Indoor Residual Spraying
*WHOPES*: World Health Organization Pesticide Evaluation Scheme
*PBO*: piperonyl butoxide
*AI*: active insecticide
*KD*: Knockdown
*LOD*: level of detection
*LOQ*: level of quantification (LOQ)
*NAFDAC*: National Agency for Food and Drug Administration and Control

## Authors’ Contributions

MOK designed the study and carried out all laboratory experiment; NOK collected the LLINs and data on net usage and treatment from users; MOK and SSI analysed and wrote the manuscript with contributions from all authors; AON and OKY offered significant insight to finalize the manuscript. All authors read, made inputs and approved the final manuscript.

## Acknowledgements

We appreciate Professor Charles Wondji for supporting and allowing us to carry out this project in his laboratory. We also thank all members of the Insectary unit of CRID for rearing the susceptible mosquitoes and assisting with the contact bioassay.

## Availability of data and materials

All data generated and analysed during this study are included in the published article.

## Ethics approval and consent to participate

Not applicable. However, verbal consent was received from the household heads after the study aim and objectives were explained to them.

## Author information

