## Supplementary material for "Contact bioassay in tandem with HPLC analysis of the active ingredients in LLINs revealed hidden possible causes of the unabated rise in malaria cases": https://drive.google.com/file/d/1vGpampD07czRTXnKvfzdH4qBWXIfqpz0/view?usp=share_link

Table S 1: List of counterfeit and fake LLINs in circulation previously reported in Nigeria

| Name of LLIN | NAFDAC Number | Remarks |
|---|---|---|
| PermaNet 2.0 | A5-0415 | NAFDAC number belongs to PermaNet 3.0 |
| Dolphin Net | None | NAFDAC Number belongs to unrelated product |
| Shielded Mosquito Net | 01-0877 | NAFDAC Number belongs to unrelated product |
| So Fine Treated Net | 01-0878 | NAFDAC Number belongs to unrelated product |
| PowerNet | 14722753 | Fake NAFDAC number |
